# Risk factor profiles of young women with vasomotor non-obstructive versus obstructive coronary syndromes: Importance of non-traditional and sex-specific risk factors

**DOI:** 10.1101/2023.05.13.23289927

**Authors:** Emilie T. Théberge, Diana N. Vikulova, Simon N. Pimstone, Liam R. Brunham, Karin H. Humphries, Tara L. Sedlak

**Affiliations:** University of British Columbia, Vancouver, BC, Canada; Centre for Heart Lung Innovation, St. Paul’s Hospital, Vancouver, BC, Canada; University of British Columbia Hospital, Vancouver, BC, Canada; Division of Cardiology, Vancouver General Hospital, Vancouver, BC, Canada

**Keywords:** Premature coronary artery disease, MINOCA, INOCA, risk factors, women

## Abstract

**Background:** Heart disease is the leading cause of premature death for women in Canada. Ischemic heart disease (IHD) is categorized as myocardial infarction (MI) with no obstructive coronary artery disease (MINOCA), ischemia with no obstructive coronary arteries (INOCA), and atherosclerotic obstructive coronary artery disease (CAD) with MI (MI-CAD) or without MI (non-MI CAD). This study aims to study the prevalence of traditional and non-traditional IHD risk factors and their relationships with (M)INOCA compared to MI-CAD and non-MI CAD in young women.

**Methods:** This study investigated women who presented with premature (≤55 years old) vasomotor entities of (M)INOCA or obstructive CAD confirmed by coronary angiography, who are currently enrolled in either the Leslie Diamond Women’s Heart Health Clinic Registry (WHC) or the Study to Avoid cardioVascular Events in BC (SAVEBC). Univariable and multivariable regression models were applied to investigate associations of risk factors with odds of (M)INOCA, MI-CAD or non-MI CAD.

**Results:** A total of 254 women enrolled between 2015-2022 were analyzed: 77 INOCA and 37 MINOCA from the WHC and 66 with non-MI CAD and 74 MI-CAD from SAVEBC. Regression analyses demonstrated that migraines and preeclampsia/gestational hypertension were the most significant risk factors with higher likelihood to associate with premature (M)INOCA relative to obstructive CAD. Conversely, the presence of diabetes and a current or previous smoking history had the highest likelihood to associate with premature CAD.

**Conclusion:** There are significant differences in the risk factor profiles of patients with premature (M)INOCA compared to obstructive CAD.

## Introduction

Ischemic heart disease (IHD) is a leading cause of preventable death of women in Canada^1^. Concerningly, over the last two decades, incidence rates have not been declining in younger women^2^. There has been a steady rise in the prevalence of several traditional modifiable cardiovascular risk factors (diabetes, hypertension, and obesity) in young women in British Columbia over the last two decades^3^, but it is unclear the degree to which these risk factors explain the lack of decline of IHD rates in young women. Moreover, there has been a growing momentum towards studying the relative contributions of “non-traditional” cardiovascular risk factors and female-specific risk factors contributing to IHD etiologies. A recent review by the Canadian Women’s Heart Health Alliance describes the higher relative risk of IHD in women compared to men who have diagnoses of risk factors such as depression, autoimmune disorders, as well as female-specific pathologies like preeclampsia^4^.

Non-obstructive IHD entities, including myocardial infarction (MI) with no obstructive coronary arteries (MINOCA) and ischemia with no obstructive coronary arteries (INOCA), are heterogenous syndromes of IHD characterized by normal or unobstructed epicardial vessels (<50% stenosis in any epicardial artery). MINOCA accounts for approximately 6% of patients presenting with ACS and is three times more prevalent in women presenting with MI (10.5%) than men (3.5%)^5^. Up to two thirds of angiograms performed for women with suspected cardiac ischemia are INOCA, which is twice as high as is observed in men^6^. Women who suffer a (M)INOCA event do not follow a benign clinical course: a study of British Columbians undergoing coronary angiography between 1999-2002 showed that the risk of major adverse cardiovascular events (MACE) in women with stable angina and nonobstructive coronary artery disease (CAD) was almost three times higher than men with nonobstructive CAD in the first year following coronary angiography^7^.

Underlying vasomotor etiologies of (M)INOCA consist predominantly of coronary vasospasm and coronary microvascular dysfunction (CMD)^8^, two entities that affect the epicardial arteries (>400µm) and microvasculature (100-400µM pre-arterioles, <100µM arterioles, and <10µm capillaries), respectively. Prevalence estimates of vasospasm and CMD are lacking in a population of women with premature IHD: CMD was identified through invasive coronary reactivity testing in approximately 50% of women with INOCA in the Women’s Ischemia Syndrome Evaluation (WISE) multicentre prospective study, which followed over 900 clinically stable women (all ages) referred for coronary angiography between 1996-2006^9^. A study by Sara et al.^10^ identified vasospasm as the responsible etiology for approximately 30% of patients who had nonobstructive CAD undergoing coronary functional testing at the Mayo Clinic catheterization lab between 1993-2012. These entities remain vastly understudied, underrecognized and undertreated compared to obstructive CAD: This paucity in recognition may be attributed to the heterogeneity of the entities, lack of robust clinical trials^11^, less knowledge about these conditions by practitioners, and difficulty obtaining specialized testing to make these diagnoses^12^.

Studies have shown that women presenting with premature acute MI (<55 years old) have differences in risk factor profiles, treatment patterns and excess mortality rates than similarly aged men^3^ and older women^13^. This may be in part due to a higher prevalence and/or differing effect sizes of traditional and non-traditional cardiovascular risk factors in this group such as higher prevalence and associated risk of MI in younger women with diabetes compared to men or their older counterparts^14^. For example, sex differences have been observed in the prevalence of major cardiovascular risk factors such as hypertension, dyslipidemia, diabetes, or smoking, and other risk enhancers^15–17^. Furthermore, Safdar et al.^18^ (2018) found fewer traditional risk factors in premature MINOCA women compared to women with MI and obstructive CAD (“MI CAD”) in the VIRGO study. It remains unclear from these studies the extent to which similar conclusions may also extend to women with INOCA and CAD with no MI (“non-MI CAD”).

Given the paucity of data on (M)INOCA in young women and the respective contributing risk factors, we sought to compare the prevalence of traditional and non-traditional risk factors in women with premature presentations of (M)INOCA and obstructive CAD, with or without MI. We specifically chose to focus on vasomotor etiologies of (M)INOCA given the heterogeneity of these entities and the prior extensive work done on other etiologies of (M)INOCA, i.e., spontaneous coronary artery dissection (SCAD)^19, 20^.

## Methods

### Study Population

This study included women with (M)INOCA enrolled in the Leslie Diamond Women’s Heart Health Clinic (WHC) Registry and women with obstructive CAD in the Study to Avoid cardioVascular Events in BC (SAVE BC) biobank.

Eligibility criteria for this study were met if women from either study had signs and symptoms of ischemia and/or an acute coronary syndrome and underwent coronary angiography (CA) and/or cardiac CT angiography (CTA) before or at 55 years old to delineate their coronary anatomy. The earliest date of CA/CTA was February 1995 for WHC and September 2015 for SAVEBC; the last date for both cohorts was July 10, 2022.

For WHC, women with MINOCA or INOCA and final diagnoses of definite or probable coronary vasospasm or CMD were included. MINOCA was defined per the 2019 Scientific Statement from the American Heart Association^21^, and INOCA was defined per the 2020 European Society of Percutaneous Cardiovascular Interventions (EAPCI) Expert Consensus Document on Ischemia with nonobstructive coronary arteries^22^. Probable and confirmed CMD and coronary vasospasm were defined using the Coronary Vasomotion Disorder International Study (COVADIS) Group definition^23^. WHC patients were excluded if their diagnosis after the first CA/CTA was obstructive CAD, non-(M)INOCA cardiac (or non-cardiac) entities (i.e., arrythmia, esophageal spasm), or if the final diagnoses were those other than vasomotor etiologies (i.e., SCAD). For SAVEBC, female patients with obstructive CAD (defined as any epicardial vessel with ≥50% stenosis) were included in this study. SAVEBC patients were excluded if there was any non-atherosclerotic component to their presentation (i.e., SCAD, vasospasm).

The WHC is a quaternary outpatient cardiology clinic located at Vancouver General Hospital (VGH) in British Columbia, Canada, comprising a multidisciplinary team of cardiologists, nurse practitioners and a psychiatrist specializing in women’s heart health. The clinic has access to specialized testing for (M)INOCA including coronary reactivity testing, optical coherence tomography (OCT), and adenosine cardiac magnetic resonance imaging (MRI). The WHC registry began recruitment of patients referred to the clinic in 2016 and has 322 patients enrolled, of which 259 (80%) patients have (M)INOCA. Data are collected from physician consult notes and diagnostic test reports uploaded to electronic medical records, including information about comorbidities (including cardiovascular risk factors) and cardiac presentation characteristics.

The SAVE BC biobank began recruitment in 2015 and is a prospective study of patients who present with premature CAD with stenosis of at least 50% in at least one epicardial artery at age ≤50 years for males and ≤55 years for females. A detailed description of the study design and rationale is provided elsewhere^24^. Briefly, and in contrast to the WHC, patients in SAVE BC were recruited from cardiac catheterization laboratories and cardiology wards at VGH, St. Paul’s Hospital (Vancouver, BC) and Kelowna General Hospital (Kelowna, BC). The SAVE BC study also collects information on cardiovascular risk factors, comorbidities, presentation characteristics, physical examination, and laboratory test results. This information is collected directly from patients during the study visits and from paper-based and electronic medical records, including medical charts, pharmacy dispensing records, and reports collected from the Cardiac Services BC Registry. The Cardiac Services BC Registry is a province-wide electronic information system collecting information on all patients who have received cardiac procedures (coronary angiography, percutaneous coronary interventions, coronary artery bypass surgery, valve procedures, and implantable devices) in the province.

This collaborative study was approved under UBC CREB application #H22-01671.

### Patient Data

Shared demographic and clinical covariates that are collected in both cohorts were identified then extracted. Variable names and definitions were standardized prior to merging the two datasets.

Sociodemographic variables were consolidated between patient self-report on study questionnaires and any mention in physician consult notes. Self-reported race (Caucasian, South Asian, East Asian, First Nations, African-Canadian, and Other) was summarized as Caucasian or non-Caucasian due to the high proportions of Caucasians in both cohorts. Partnered status was recorded as partnered (married or common law) or not partnered (single, divorced, or widowed).

Traditional cardiovascular risk factors included: dyslipidemia (total cholesterol ≥240 mg/dL (6.2 mmol/L), low-density lipoprotein cholesterol (LDL-C) ≥160 mg/dL (4.1 mmol/L), high density lipoprotein cholesterol (HDL-C) ≤ 40 mg/dL (1.0 mmol/L), triglycerides (TG) ≥200 mg/dL (2.3 mmol/L), or treatment of dyslipidemia^25^), hypertension (systolic/diastolic blood pressure (SBP/DBP) ≥140/90 mmHg as per Canadian Hypertension Guidelines^26^), diabetes (type 1 or 2: fasting plasma glucose ≥126 mg/dL (7 mmol/L) in at least 2 baseline measurements, hemoglobin A1c ≥ 6.5%, physician diagnosis, or treatment of diabetes^27^), family history of premature cardiovascular disease (CVD) (<55 years old for men, <65 for women), obesity (BMI ≥30 kg/m^2^), and smoking (former, current, or never).

Non-traditional clinical risk factors and female-specific risk modifiers included: history of a clinical diagnosis of depression and/or anxiety, hypothyroidism, diagnosis of migraine (with or without aura), diagnosis of an autoimmune disease (AID), preeclampsia (gestational SBP/DBP ≥140/90 mmHg and proteinuria per American College of Obstetricians and Gynecologists (ACOG) guidelines^28^) or gestational hypertension, gestational diabetes, number of children (0-1, 2-3, 4+), and number of non-term pregnancies such as miscarriage, abortion and/or stillbirth (0, 1, or 2+).

### Data Analyses

Baseline characteristics were reported as medians for continuous variables and counts with proportions for categorical variables. Cumulative burden was calculated by summing the number of traditional CV risk factors (range 0-6) and non-traditional CV risk factors (range 0-5). The main comparison was between the two cohorts; both cohorts were further stratified by the presence/absence of MI. Categorical variables were compared using Chi-squared tests; continuous variables were compared using the Kruskal-Wallis test.

To assess the strength of the association between each risk factor and the relevant diagnoses, several univariable logistic regression tests were conducted. The primary comparisons of interest were associations with (M)INOCA relative to any obstructive CAD. The associations between each risk factor and diagnoses were then assessed for INOCA relative to non-MI CAD, and MINOCA relative to MI-CAD, with the associations expressed as odds.

Iterative, multivariable logistic regression models were constructed to identify factors independently associated with (M)INOCA compared to any obstructive CAD. In the first model, covariates were selected based on clinical importance: age at earliest angiography; diabetes (type 1 (T1) or type 2 (T2)); hypertension; depression/anxiety; and family history of premature CVD. In the second model, the three variables with the strongest effect sizes in the univariable analysis were added to the model: preeclampsia/gestational hypertension, smoking status and migraines. Log likelihood changes, Akaike’s criterion and number of degrees of freedom were assessed to optimize the model fit.

## Results

### Baseline characteristics

There were 114 women with premature (M)INOCA from a vasomotor entity from the WHC cohort (n=37 MINOCA, n=77 INOCA) and 140 women with premature obstructive CAD from the SAVEBC cohort (n=74 MI-CAD, n=66 non-MI CAD) that met inclusion criteria, as shown in Figure 1.

**Figure 1.**
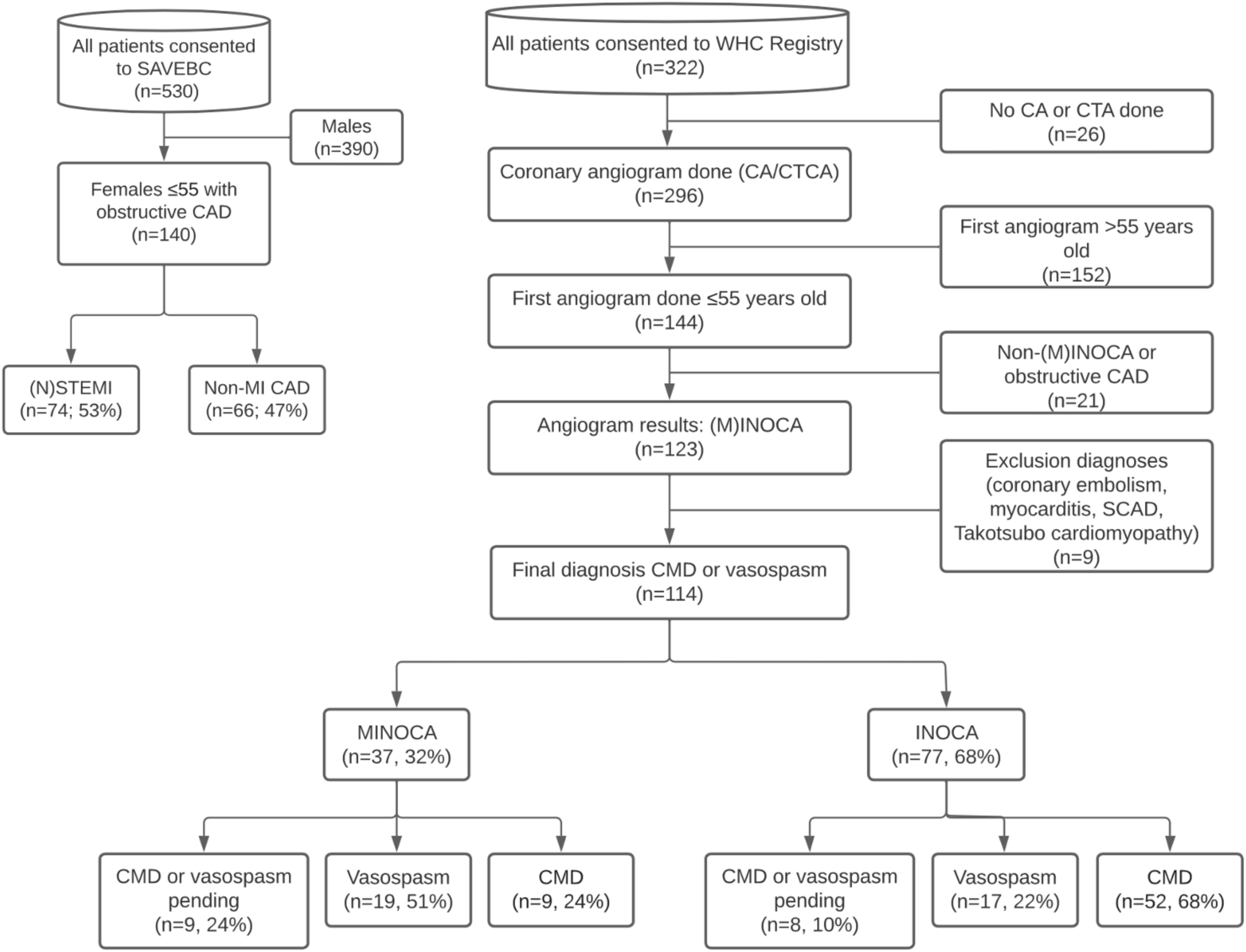
Flow diagram of exclusion steps by which eligible SAVEBC and WHC patients were filtered.

In the WHC cohort, most women with MINOCA had probable or definite vasospasm (19, 51%), the rest had probable or definitive CMD (9, 24%) or an undetermined diagnosis of probable CMD or vasospasm still awaiting final testing, but with non-vasomotor entities ruled out (9, 24%). Most women with INOCA had CMD (52, 68%), and the rest had vasospasm (17, 22%) and an undetermined diagnosis of CMD or vasospasm (8, 10%). In the SAVEBC cohort, 50 women with MI-CAD presented with NSTEMI (68%) and the other 24 with STEMI (32%). Among the non-MI CAD, 35 presented with stable angina (53%), 22 with unstable angina (33%), and 9 with other indications for undergoing coronary angiography (14%).

### Risk factors

Table 1 summarizes the frequency of demographic and clinical variables across diagnostic subgroups. The median ages (IQR) at the first coronary angiography in each subgroup of women were 49 years (42,55) for MINOCA, 50 (41,55) for INOCA, 50 (44,55) for MI-CAD, and 50.5 (44.5,55) for non-MI CAD. Significant differences were found between women with (M)INOCA (WHC) compared to obstructive CAD (SAVEBC): (M)INOCA women had higher proportions of self-reported Caucasian race (p<0.01), being partnered (p<0.01), history of migraine (p<0.001), of preeclampsia or gestational hypertension (p=0.04), and more children. In contrast, women with obstructive CAD had significantly higher proportions of dyslipidemia (p<0.001), diabetes (p<0.001), hypertension (p<0.01), obesity (p<0.001) and smoking history (p<0.001).

**Table 1.** Frequencies of risk factors in the WHC and SAVEBC cohorts further stratified by diagnostic subgroups (MINOCA, INOCA, MI-CAD, non-MI CAD).

|  | WHC |  | SAVEBC |  | WHC<br>(n=114) | SAVEBC<br>(n=140) | p-value<br>(WHC<br>vs.<br>SAVEBC) |
| --- | --- | --- | --- | --- | --- | --- | --- |
|  | ≤55<br>MINOCA<br>(n=37) | ≤55<br>INOCA<br>(n=77) | ≤55 MI-CAD (n=74) | ≤55 non-MI<br>CAD (n=66) |  |  |  |
| **Age at first CA/CTA** (years, IQR) | 49 (42,55) | 50 (41,55) | 50 (44,55) | 50.5 (44.5,55) | 49 (41,55) | 50 (44,55) | 0.08 |
| **Race, n (%)** |  |  |  |  |  |  |  |
| **Caucasian** (“White”) | 24 (65) | 56 (73) | 30 (41) | 27 (41) | 80 (70) | 57 (41) | <0.01 |
| **Non-Caucasian** | 11 (30) | 20 (26) | 29 (39) | 25 (38) | 31 (27) | 54 (39) |  |
| South Asian | 2 (5) | 7 (9) | 7 (9) | 5 (8) | 9 (8) | 12 (9) |  |
| East Asian (“Chinese, Japanese, Korean”) | 5 (14) | 2 (3) | 5 (7) | 4 (6) | 7 (6) | 9 (6) |  |
| First Nations (“Aboriginal”) | 0 | 2 (3) | 6 (8) | 2 (3) | 2 (2) | 8 (6) |  |
| African-Canadian (“Black”) | 0 | 1 (1) | 0 | 0 | 1 (1) | 0 |  |
| Other (incl. Mixed) | 4 (11) | 8 (10) | 11 (15) | 14 (21) | 12 (11) | 25 (18) |  |
| *Missing\** | 2 (5) | 1 (1) | 15 (20) | 14 (21) | 3 (3) | 29 (21) |  |
| **Partnered** (Married, common law), n (%) | 36 (97) | 61 (79) | 36 (49) | 32 (48) | 97 (85) | 68 (49) | <0.01 |
| *Missing, n* | 0 | 0 | 18 (24) | 20 (30) | 0 | 38 (27) |  |
| **Dyslipidemia, n (%)** | 14 (38) | 39 (51) | 46 (62) | 48 (73) | 53 (46) | 94 (67) | <0.001 |
| **Diabetes, n (%)** | 3 (8) | 10 (13) | 24 (32) | 22 (33) | 13 (11) | 46 (33) | <0.001 |
| Type 1 | 0 | 2 (3) | 0 | 2 (3) | 2 (2) | 2 (1) |  |
| Type 2 | 3 (8) | 8 (10) | 24 (32) | 20 (30) | 11 (10) | 44 (31) |  |
| Gestational Diabetes | 3 (8) | 8 (10) | 9 (12) | 16 (24) | 11 (10) | 25 (18) |  |
| **Hypertension, n (%)** | 13 (35) | 30 (39) | 42 (57) | 34 (52) | 43 (38) | 76 (54) | <0.01 |
| **Obesity (BMI ≥30), n (%)** | 8 (22) | 17 (22) | 35 (47) | 26 (39) | 25 (22) | 61 (44) | <0.001 |
| *Missing* | 0 | 0 | 5 (7) | 1 (2) | 0 | 6 (4) |  |
| **Smoking history, n (%)** |  |  |  |  |  |  |  |
| Never | 33 (89) | 66 (86) | 34 (46) | 38 (58) | 99 (87) | 72 (51) | <0.001 |
| Current | 1 (3) | 2 (3) | 17 (23) | 9 (14) | 3 (3) | 26 (19) |  |
| Previous | 3 (8) | 9 (12) | 23 (31) | 19 (29) | 12 (11) | 42 (30) |  |
| **Family history of premature CVD, n (%)** | 16 (43) | 31 (40) | 31 (42) | 32 (48) | 47 (41) | 63 (45) | 0.1 |
| *Missing* | 0 | 0 | 15 (20) | 5 (8) | 0 | 20 (14) |  |
| **Depression/Anxiety, n (%)** | 13 (35) | 26 (34) | 22 (30) | 19 (29) | 39 (34) | 41 (29) | 0.54 |
| **Hypothyroidism, n (%)** | 2 (5) | 14 (18) | 12 (16) | 10 (15) | 16 (14) | 22 (16) | 0.6 |
| **Migraines, n (%)** | 19 (51) | 23 (30) | 2 (3) | 5 (8) | 42 (37) | 7 (5) | <0.001 |
| **Autoimmune Disorder(s) (any), n (%)** | 3 (8) | 14 (18) | 18 (24) | 12 (18) | 17 (15) | 30 (21) | 0.21 |
| **Preeclampsia or gestational hypertension, n (%)** | 7 (19) | 6 (8) | 3 (4) | 2 (3) | 13 (11) | 5 (4) | 0.04 |
| *Missing* | 4 (11) | 10 (13) | 19 (26) | 15 (23) | 14 (12) | 34 (24) |  |
| **Number of children, n (%)** |  |  |  |  |  |  |  |
| 0 or 1 | 8 (22) | 12 (16) | 21 (28) | 21 (32) | 20 (18) | 42 (30) | 0.04 |
| 2 or 3 | 22 (59) | 50 (65) | 36 (49) | 31 (47) | 72 (63) | 67 (48) |  |
| 4+ | 3 (8) | 5 (6) | 6 (8) | 3 (5) | 8 (7) | 9 (6) |  |
| *Missing* | 4 (11) | 10 (13) | 11 (15) | 11 (17) | 14 (12) | 22 (16) |  |
| **Number of non-term pregnancies, n (%)** |  |  |  |  |  |  |  |
| 0 | 18 (49) | 25 (32) | 32 (43) | 33 (50) | 43 (38) | 65 (46) | 0.68 |
| 1 | 7 (19) | 16 (21) | 16 (22) | 14 (21) | 23 (20) | 30 (21) |  |
| 2+ | 2 (5) | 18 (23) | 14 (19) | 8 (12) | 20 (18) | 22 (16) |  |
| *Missing* | 10 (27) | 18 (23) | 12 (16) | 11 (17) | 28 (25) | 23 (16) |  |

\* *Missing proportions provided when greater than 5%.*

The cumulative burden of traditional and non-traditional risk factors is summarized in Tables 2 and 3, respectively, with relative proportions visualized in Figure 2 (traditional risk factors) and Figure 3 (non-traditional risk factors). Chi-squared tests comparing cumulative burden of risk factors demonstrate significant differences between cohorts in the number of traditional risk factors (Figure 2A, p<0.001) and non-traditional risk factors (Figure 3A, p=0.03). The most frequent number of traditional risk factors seen per patient was higher for the patients with obstructive CAD (mode=3), compared women with non-obstructive CAD (mode=1). Most strikingly, over 40% of the MINOCA group had just one traditional risk factor. The opposite trend was seen with non-traditional risk factors: in women with non-obstructive CAD the most common number of risk factors was one; in women with obstructive CAD the most common number was zero.

**Figure 2.**
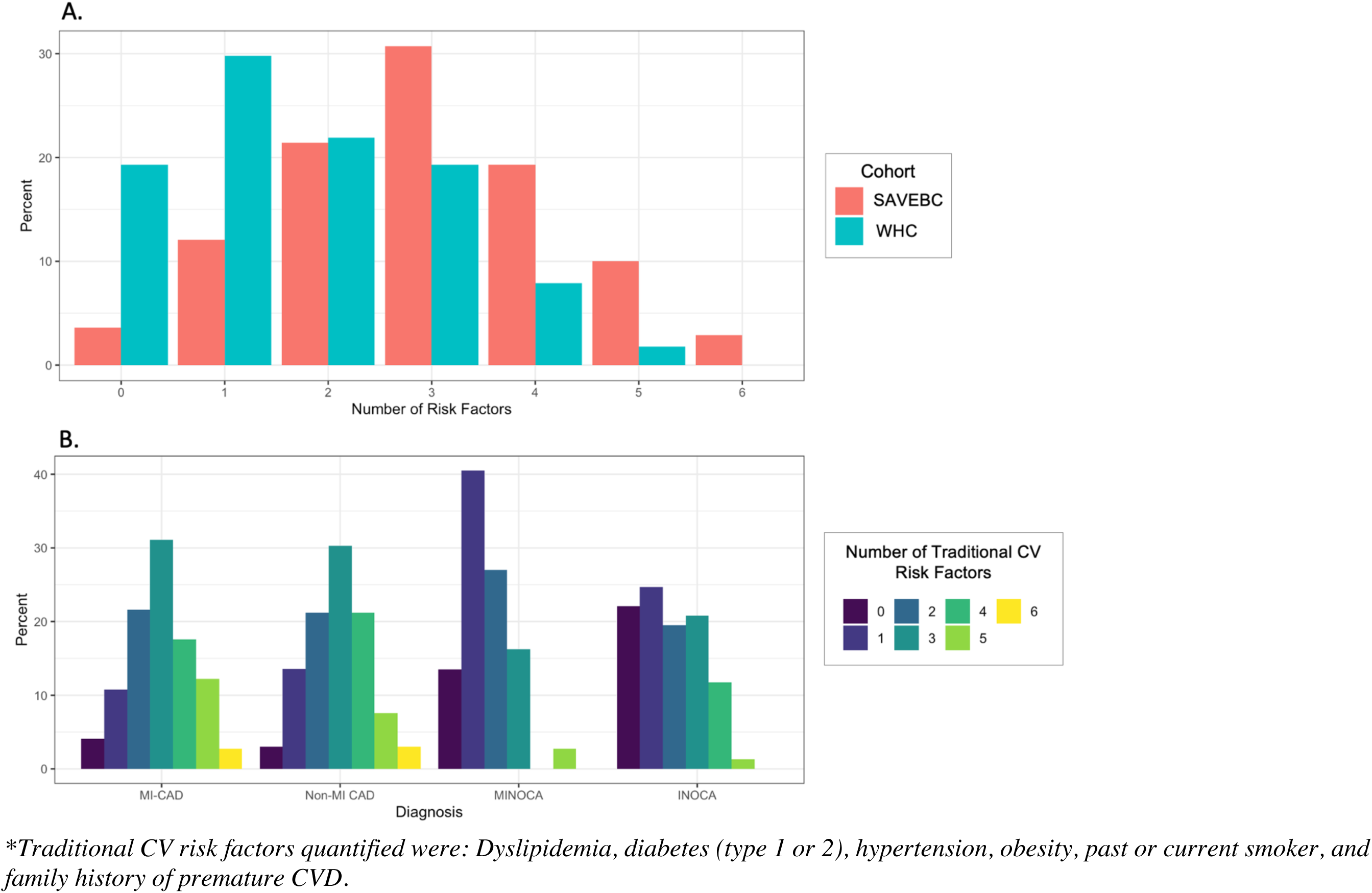
Histograms of total traditional CV risk factors* by: **A.** Cohort and **B.** Diagnostic Subgroup (MI-CAD vs non-MI CAD vs MINOCA vs INOCA).

**Figure 3.**
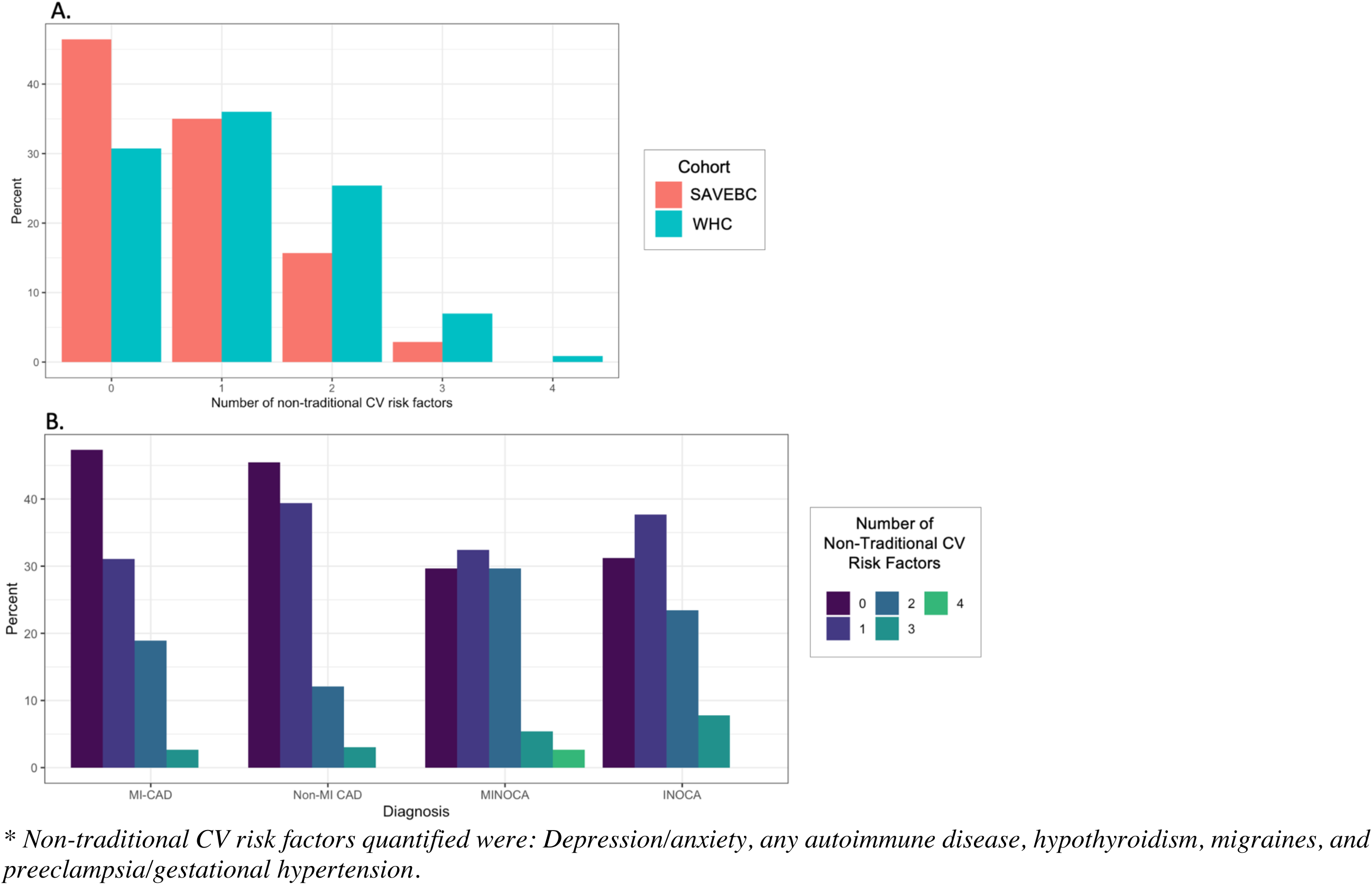
Histograms of total non-traditional CV risk factors by: **A.** Cohort and **B.** Diagnostic Subgroup (MI-CAD vs non-MI CAD vs MINOCA vs INOCA).

**Table 2.**
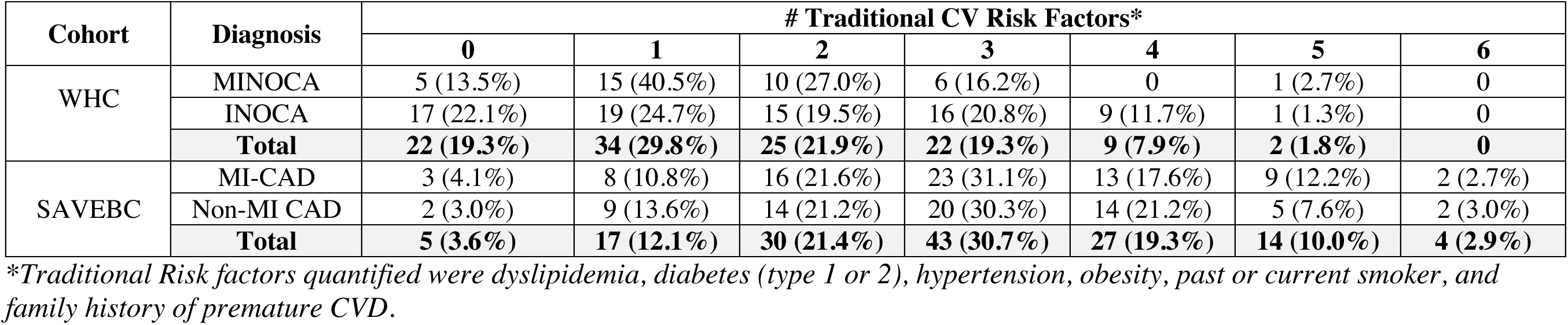
Traditional CV risk factors stratified by cohort (SAVEBC vs WHC) and by diagnostic subgroup (MI-CAD, non-MI CAD, MINOCA, INOCA).

| Cohort | Diagnosis | # Traditional CV Risk Factors* |  |  |  |  |  |  |
| --- | --- | --- | --- | --- | --- | --- | --- | --- |
|  |  | 0 | 1 | 2 | 3 | 4 | 5 | 6 |
| WHC | MINOCA | 5 (13.5%) | 15 (40.5%) | 10 (27.0%) | 6 (16.2%) | 0 | 1 (2.7%) | 0 |
|  | INOCA | 17 (22.1%) | 19 (24.7%) | 15 (19.5%) | 16 (20.8%) | 9 (11.7%) | 1 (1.3%) | 0 |
|  | **Total** | **22 (19.3%)** | **34 (29.8%)** | **25 (21.9%)** | **22 (19.3%)** | **9 (7.9%)** | **2 (1.8%)** | **0** |
| SAVEBC | MI-CAD | 3 (4.1%) | 8 (10.8%) | 16 (21.6%) | 23 (31.1%) | 13 (17.6%) | 9 (12.2%) | 2 (2.7%) |
|  | Non-MI CAD | 2 (3.0%) | 9 (13.6%) | 14 (21.2%) | 20 (30.3%) | 14 (21.2%) | 5 (7.6%) | 2 (3.0%) |
|  | **Total** | **5 (3.6%)** | **17 (12.1%)** | **30 (21.4%)** | **43 (30.7%)** | **27 (19.3%)** | **14 (10.0%)** | **4 (2.9%)** |

*\*Traditional Risk factors quantified were dyslipidemia, diabetes (type 1 or 2), hypertension, obesity, past or current smoker, and family history of premature CVD.*

**Table 3.** Non-traditional CV risk factors stratified by cohort (SAVEBC vs WHC) and by diagnostic subgroup (MI-CAD, non-MI CAD, MINOCA, INOCA).

| Cohort | Diagnosis | # Non-Traditional CV Risk Factors* |  |  |  |  |  |
| --- | --- | --- | --- | --- | --- | --- | --- |
|  |  | 0 | 1 | 2 | 3 | 4 | 5 |
| WHC | MINOCA | 11 (29.7%) | 12 (32.4%) | 11 (29.7%) | 2 (5.4%) | 1 (2.7%) | 0 |
|  | INOCA | 24 (31.2%) | 29 (37.7%) | 18 (25.4%) | 6 (7.8%) | 0 | 0 |
|  | **Total** | **35 (30.7%)** | **41 (36.0%)** | **29 (25.4%)** | **8 (7.0%)** | **1 (0.9%)** | **0** |
| SAVEBC | MI-CAD | 35 (47.3%) | 23 (31.1%) | 14 (18.9%) | 2 (2.7%) | 0 | 0 |
|  | Non-MI CAD | 30 (45.5%) | 26 (39.4%) | 8 (12.1%) | 2 (3.0%) | 0 | 0 |
|  | **Total** | **65 (46.4%)** | **49 (35.0%)** | **22 (15.7%)** | **4 (2.9%)** | **0** | **0** |

*\*Non-traditional CV risk factors quantified were depression/anxiety, any autoimmune disease, hypothyroidism, migraines, and preeclampsia/gestational hypertension.*

Among the 30% of WHC women with only one traditional risk factor, family history of premature CVD was the most common (38%), followed by dyslipidemia (24%) then hypertension and obesity (15% each). Among the 12% of SAVEBC patients with only one traditional risk factor, the most common were dyslipidemia (29%) and smoking history (29%), followed by family history of premature CVD (18%). Having 5 or 6 traditional CV risk factors was observed in over six times more patients in the SAVEBC cohort (12.9%) compared to the WHC (1.8%).

For non-traditional cardiovascular risk factors, the most frequently reported number of risk factors among WHC patients was one (36%). The most common was a history of migraine (37%), followed by depression/anxiety (34%) and hypothyroidism (12%). Among the SAVEBC patients, 46.5% had zero non-traditional risk factors, but among the 35% with only one non traditional risk factor, the most common was depression/anxiety (45%), followed by any autoimmune disease (31%) and hypothyroidism (18%). Having 3 or 4 non-traditional risk CV risk factors was observed over twice as often in the WHC patients (7.9%) compared to SAVEBC patients (2.9%)

### Univariable Logistic Regression Models

Odds ratios (OR) and 95% confidence intervals (CI) from the univariable regression models are summarized in Table 4. Most traditional risk factors, except for family history of CVD, were significantly associated with a reduced odds of (M)INOCA compared to any obstructive CAD, shown in Figure 4A. The risk factors most strongly associated with an increased odds of (M)INOCA (OR>2) were history of migraines OR 10.50 (4.75,26.66), p<0.001; preeclampsia or gestational hypertension OR 3.02 (1.09,9.72), p=0.04; and having 2-3 children OR 2.26 (1.22,4.29), p=0.011.

**Figure 4.**
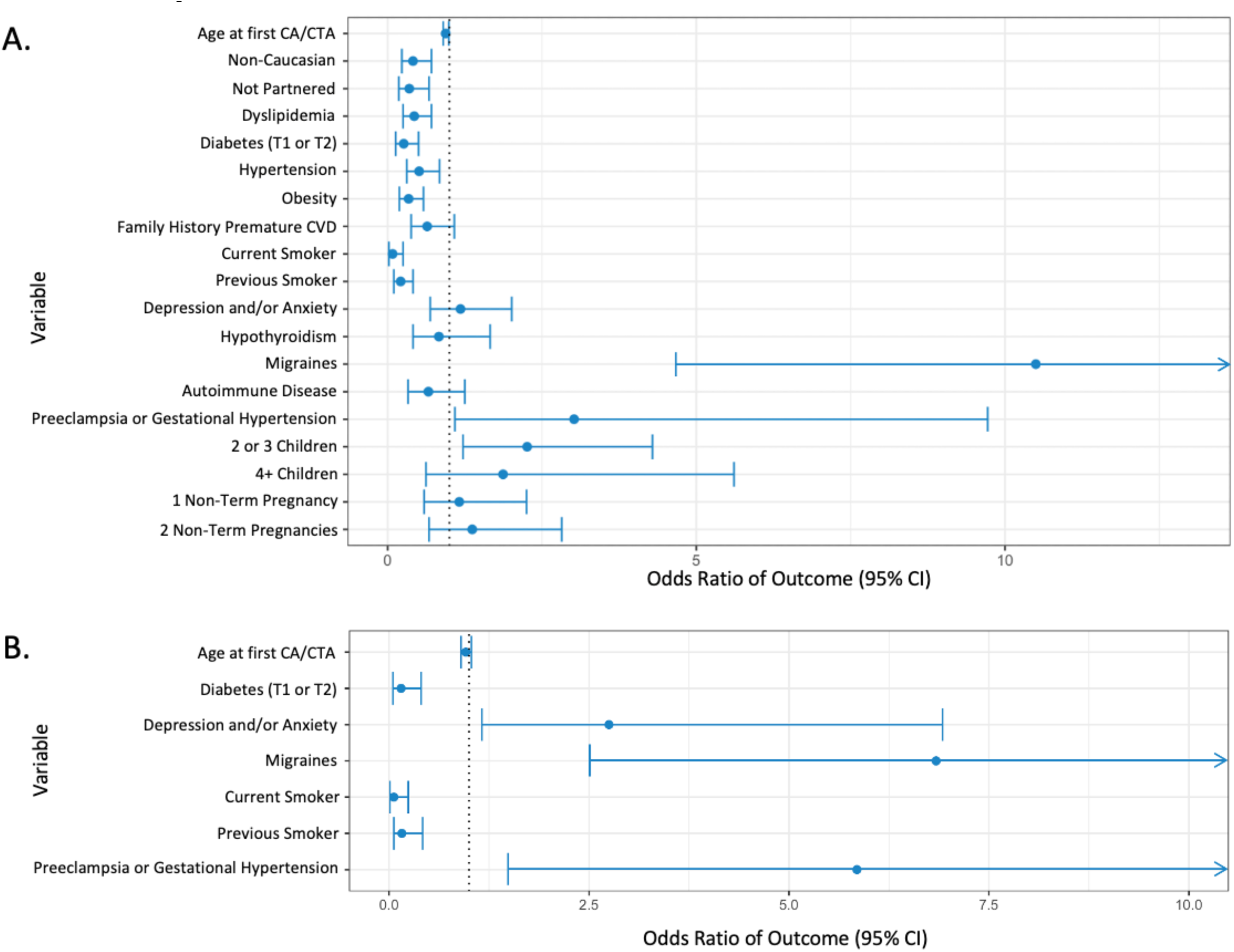
Forest plots of odds ratios with 95% CI for (M)INOCA compared to any obstructive CAD. **A.** Univariable odds ratios, **B.** Multivariable (adjusted) odds ratios.

**Table 4.** Association between risk factors and selected diagnoses. Odds ratio (OR) and 95% confidence interval (CI) from univariable logistic regression models.

| Outcome |  | (M)INOCA<br>(Referent: any obstructive CAD*)<br>Odds ratio<br>(95% CI) | INOCA<br>(Referent: non-MI CAD)<br>Odds ratio<br>(95% CI) | MINOCA<br>(Referent: MI-CAD)<br>Odds ratio<br>(95% CI) |
| --- | --- | --- | --- | --- |
| Sociodemographic Variables | Age at earliest CA/CTA (per year) | **0.94 (0.90,0.99)** | 0.95 (0.89,1.00) | 0.94 (0.87,1.00) |
|  | Race: Non-Caucasian | **0.41 (0.23,0.71)** | **0.39 (0.18,0.81)** | 0.47 (0.19,1.12) |
|  | Not partnered | **0.35 (0.18,0.67)** | 0.60 (0.26,1.38) | **0.05 (0.00,0.26)** |
| Traditional CV Risk Factors | Dyslipidemia | **0.43 (0.25,0.71)** | **0.38 (0.19,0.78)** | **0.37 (0.16,0.83)** |
|  | Diabetes | **0.26 (0.13,0.50)** | **0.30 (0.13,0.69)** | **0.18 (0.04,0.58)** |
|  | Hypertension | **0.51 (0.31,0.84)** | 0.60 (0.31,1.17) | **0.41 (0.18,0.92)** |
|  | Obese (BMI >=30) | **0.34 (0.19,0.58)** | **0.43 (0.20,0.88)** | **0.27 (0.10,0.65)** |
|  | Family history of premature CVD | 0.64 (0.38,1.08) | 0.62 (0.32,1.23) | 0.69 (0.30,1.57) |
|  | Smoking history: Current | **0.08 (0.02,0.25)** | **0.13 (0.03,0.62)** | **0.06 (0.00,0.32)** |
|  | Smoking history: Former | **0.21 (0.10,0.41)** | **0.27 (0.11,0.66)** | **0.13 (0.03,0.43)** |
| Non-traditional Risk Factors & Female-specific Variables | Depression/Anxiety | 1.18 (0.69,2.01) | 1.21 (0.59,2.47) | 1.18 (0.50,2.73) |
|  | Hypothyroidism | 0.83 (0.41,1.66) | 1.20 (0.49,2.92) | 0.28 (0.04,1.09) |
|  | Migraines | **10.50 (4.75,26.66)** | **4.94 (1.75,13.92)** | **36.89 (9.30,238.93)** |
|  | Any Autoimmune Disorder(s) | 0.66 (0.33,1.25) | 1.02 (0.43,2.38) | 0.28 (0.06,0.92) |
|  | Preeclampsia/Gestational Hypertension | **3.02 (1.09,9.72)** | 2.41 (0.47,12.47) | **4.67 (1.19,23.05)** |
|  | Number of children: 2 to 3 | **2.26 (1.22,4.29)** | **2.82 (1.22,6.53)** | 1.60 (0.62,4.42) |
|  | Number of children: 4+ | 1.87 (0.62,5.61) | 2.92 (0.59,14.41) | 1.31 (0.23,6.37) |
|  | Number of non-term pregnancies: 1 | 1.16 (0.59,2.25) | 1.51 (0.62,3.66) | 0.78 (0.26,2.20) |
|  | Number of non-term pregnancies: 2+ | 1.37 (0.67,2.82) | **2.97 (1.11,7.93)** | 0.25 (0.04,1.05) |

\* Any obstructive CAD: includes MI-CAD and non-MI CAD

Risk factors known to be strongly associated with obstructive CAD were strongly and negatively associated with (M)INOCA (OR<0.5): a current smoker OR 0.08 (0.02,0.25) p<0.001; former smoker OR 0.21 (0.10,0.41, p<0.001; T1 or T2 diabetes OR 0.26 (0.13,0.50), p<0.001; obesity OR 0.34 (0.19,0.58), p<0.001; not partnered OR 0.35 (0.18,0.67), p=0.002; non-Caucasian race OR 0.41 (0.23,0.71), p=0.002; and dyslipidemia OR 0.43 (0.25,0.71), p=0.001.

Risk factors not significantly associated with (M)INOCA compared to any obstructive CAD were family history of CVD, depression/anxiety, hypothyroidism, and any autoimmune disease.

### Multivariable Regression Models

After iterative multivariable regression models were fitted and assessed, the final model of (M)INOCA relative to any obstructive CAD included (Figure 4B): age of earliest angiography OR 0.96 (0.90,1.03), p=0.25; diabetes OR 0.15 (0.05,0.40), p<0.001); depression/anxiety OR 2.75 (1.16,6.92), p=0.025; migraines OR 6.84 (2.56,21.74), p<0.001; current smoking history OR 0.06 (0.01,0.24), p<0.001; previous smoking history OR 0.16 (0.06,0.42), p<0.001; and preeclampsia or gestational hypertension OR 5.85 (1.40,30.58), p=0.022.

## Discussion

This study demonstrated distinct differences in risk factor profiles of women with premature (M)INOCA – notably non-traditional CV risk factors such as migraines, preeclampsia/gestational hypertension, depression/anxiety – compared to premature obstructive CAD, characterized mostly by traditional CV risk factors. These differences suggest different mechanisms and etiologies, underscoring a need for clinicians to collect and assess emerging non-traditional and sex-specific variables as risk factors that put patients at higher relative risk of (M)INOCA.

The results of this study support the hypothesis of possible shared vasomotor disorder mechanisms between (M)INOCA, migraines and preeclampsia/gestational hypertension. This concept of a systemic vasomotor disorder is not new; previous studies have identified strong associations with migraines and Raynaud’s syndrome in patients with vasospastic angina^29^. However, there is a paucity of molecular (i.e., genetic) studies of vasomotor (M)INOCA etiologies that could connect shared molecular mechanisms of these disorders. There are multiple proposed routes towards dysfunctional modulation of vascular tone by the endothelium: from enhanced coronary vasoconstrictive reactivity and microvascular level, impaired endothelium-independent and dependent vasodilatory capacity, to increased microvascular resistance due to structural factors (e.g., vascular remodelling, luminal narrowing)^30^. Furthermore, there may be a sex-specific effect of high estrogen levels (relative to post-menopausal levels) on subtypes of these pathologies, as estrogen promotes nitric oxide (NO) production via activation of NO synthase and subsequent endothelium-dependent vasodilation^31^. More research is required to understand the differences and shared mechanisms between cardiac vasomotor subtypes with non-cardiac vasomotor etiologies (i.e., migraines), and the role of sex hormones.

Noteworthy was the emergence of depression/anxiety as an independent variable in the adjusted multivariable model, as it was not significant in the univariable models for (M)INOCA. Depression and anxiety are highly comorbid conditions with each other and occur in women with IHD more frequently than in men with IHD^32^. A persistent depressive state has been associated with abnormal vascular reactivity, predominantly through autonomic dysfunction and cumulative effects over time from chronic low-grade inflammation^33^. Depression and anxiety can have acute and chronic effects on the dysregulation of vasovagal tone through persisting activation of the hypothalamic-pituitary-adrenal axis, which exhibits sex differences in vasoconstrictive responsiveness^34^. Thus, prolonged activation may further exacerbate risk of (M)INOCA in women with higher predisposition towards vasomotor syndromes. Altogether, these findings support a complex and bidirectional mechanism of depression/anxiety contributing to (M)INOCA risk.

Pregnancy has lifelong impacts on women’s physiology, with numerous studies demonstrating pregnancy’s long-term benefits on immune^35^ and cardiovascular health^36^. A proposed explanation for this link is improved endothelial function resulting in greater bioavailable NO during pregnancy that persists postpartum^37, 38^. In the current study, having 2-3 children was associated with increased odds of (M)INOCA, which may also be interpreted as having 2-3 children is associated with a lower risk of CAD. This relationship has been previously reported as a J-shaped curve, wherein 2-3 children is associated with fewer cardiovascular events than 0-1 or 4+ children^39^. Because our findings relate to (M)INOCA relative to CAD, this question needs to be examined in future studies comparing women with CAD or (M)INOCA to women with no IHD.

The prevalence of traditional CV risk factors, dyslipidemia, diabetes and smoking status, in our (M)INOCA cohort is similar to that of the general Canadian population of 45%, 11% and 10%, respectively^40–42^. This contrasts with migraines, which were observed over four times as frequently in the (M)INOCA cohort (36%) compared to the Canadian population (8.3%)^43^. Focusing on traditional CV risk factors allows for reasonable prediction of possible premature obstructive CAD but not premature (M)INOCA, as most (M)INOCA patients did not have these traditional risk factors.

Limitations of this study include its small sample size (particularly of MINOCA patients), lack of racial/ethnic diversity, and the omission of other sex-specific variables emerging as CV risk factors due to absence of systematic collection in either or both cohorts, such as premature menopause (<45 years old), endometriosis and polycystic ovarian syndrome. It is unclear the extent to which the results from this study would apply to non-Caucasian populations; for example, several studies have shown significantly higher prevalence of vasospasm by provocative testing in people of Japanese^44^, Taiwanese^45^ and South Korean^46^ descent compared to Caucasians^22^. However, the risk factor profiles of the young women within these populations are not clear.

In conclusion, our study on risk factors associated with vasomotor etiologies of premature (M)INOCA compared to those associated with premature obstructive CAD provides further evidence that these entities demonstrate distinct risk factor profiles in young women. Specifically, non-traditional risk factors such as migraines, preeclampsia/gestational hypertension and depression/anxiety increase the risk of (M)INOCA and should be reliably documented in all patients presenting with possible IHD. Future studies with larger sample sizes, both sexes and systematic collection of non-traditional risk factors are needed to improve our understanding of the mechanisms of vasomotor pathology in these premature-onset populations.

## Data Availability

All data produced in the present study are available upon reasonable request to the authors.

## Acknowledgments

The authors would like to thank statistician May Lee for her coding support in a portion of the statistical analyses.

## References

1. Heart and Stroke Foundation. Ms.Understood. 2018 Heart Report. Available at: https://www.heartandstroke.ca/-/media/pdf-files/canada/2018-heart-month/hs_2018-heart-report_en.ashx Accessed on December 20th, 2022.

2. Botly, L. C. P., Lindsay, M. P., Mulvagh, S. L., Hill, M. D., Goia, C., Martin-Rhee, M., Casaubon, L. K., & Yip, C. Y. Y. (2020). Recent Trends in Hospitalizations for Cardiovascular Disease, Stroke, and Vascular Cognitive Impairment in Canada. The Canadian journal of cardiology, 36(7), 1081–1090. https://doi.org/10.1016/j.cjca.2020.03.007

3. Vikulova, D. N., Grubisic, M., Zhao, Y., Lynch, K., Humphries, K. H., Pimstone, S. N., & Brunham, L. R. (2019). Premature Atherosclerotic Cardiovascular Disease: Trends in Incidence, Risk Factors, and Sex-Related Differences, 2000 to 2016. Journal of the American Heart Association, 8(14), e012178. https://doi.org/10.1161/JAHA.119.012178

4. Mulvagh, S. L., Mullen, K. A., Nerenberg, K. A., Kirkham, A. A., Green, C. R., Dhukai, A. R., Grewal, J., Hardy, M., Harvey, P. J., Ahmed, S. B., Hart, D., Levinsson, A. L. E., Parry, M., Foulds, H. J. A., Pacheco, C., Dumanski, S. M., Smith, G., & Norris, C. M. (2021). The Canadian Women’s Heart Health Alliance Atlas on the Epidemiology, Diagnosis, and Management of Cardiovascular Disease in Women – Chapter 4: Sex and Gender-Unique Disparities: CVD Across the Lifespan of a Woman. CJC open, 4(2), 115–132. https://doi.org/10.1016/j.cjco.2021.09.013

5. Mahajan, A. M., Gandhi, H., Smilowitz, N. R., Roe, M. T., Hellkamp, A. S., Chiswell, K., Gulati, M., & Reynolds, H. R. (2019). Seasonal and circadian patterns of myocardial infarction by coronary artery disease status and sex in the ACTION Registry-GWTG. International Journal of Cardiology, 274, 16–20. https://doi.org/10.1016/j.ijcard.2018.08.103

6. Bairey Merz, C. N., Pepine, C. J., Walsh, M. N., & Fleg, J. L. (2017). Ischemia and No Obstructive Coronary Artery Disease (INOCA): Developing Evidence-Based Therapies and Research Agenda for the Next Decade. Circulation, 135(11), 1075–1092. https://doi.org/10.1161/CIRCULATIONAHA.116.024534

7. Sedlak, T. L., Lee, M., Izadnegahdar, M., Merz, C. N., Gao, M., & Humphries, K. H. (2013). Sex differences in clinical outcomes in patients with stable angina and no obstructive coronary artery disease. American Heart Journal, 166(1), 38–44. https://doi.org/10.1016/j.ahj.2013.03.015

8. Sinha, A., Rahman, H., & Perera, D. (2020). Coronary microvascular disease: current concepts of pathophysiology, diagnosis and management. Cardiovascular endocrinology & metabolism, 10(1), 22–30. https://doi.org/10.1097/XCE.0000000000000223

9. Reis, S. E., Holubkov, R., Conrad Smith, A. J., Kelsey, S. F., Sharaf, B. L., Reichek, N., Rogers, W. J., Merz, C. N., Sopko, G., Pepine, C. J., & WISE Investigators (2001). Coronary microvascular dysfunction is highly prevalent in women with chest pain in the absence of coronary artery disease: results from the NHLBI WISE study. American heart journal, 141(5), 735–741. https://doi.org/10.1067/mhj.2001.114198

10. Sara, J. D., Widmer, R. J., Matsuzawa, Y., Lennon, R. J., Lerman, L. O., & Lerman, A. (2015). Prevalence of Coronary Microvascular Dysfunction Among Patients With Chest Pain and Nonobstructive Coronary Artery Disease. JACC. Cardiovascular interventions, 8(11), 1445– 1453. https://doi.org/10.1016/j.jcin.2015.06.017

11. Handberg, E., Merz, C. N. B., Cooper-Dehoff, R.M., Wei, J., Conlon, M., Lo, M.C., Boden, W., Frayne, S.M., Villines, T., Spertus, J.A., Weintraub, W., O’Malley, P., Chaitman, B., Shaw, L.J., Budoff, M., Rogatko, A., & Pepine, C.J. (2021). Rationale and design of the Women’s Ischemia Trial to Reduce Events in Nonobstructive CAD (WARRIOR) trial. The American Heart Journal, 237, 90–103. https://doi.org/10.1016/j.ahj.2021.03.011

12. Pacheco, C., Mullen, K.A., Coutinho, T., Jaffer, S., Parry, M., Van Spall, H., Clavel, M.A., Edwards, J.D., Sedlak, T.L., Norris, C.M., Dhukai, A., Grewal, J., & Mulvagh, S.L. (2021). The Canadian Women’s Heart Health Alliance Atlas on the Epidemiology, Diagnosis, and Management of Cardiovascular Disease in Women – Chapter 5: Sex and Gender-Unique Manifestations of Cardiovascular Disease. CJC open, 4(3), 243–262. https://doi.org/10.1016/j.cjco.2021.11.006

13. Lichtman, J. H., Lorenze, N. P., D’Onofrio, G., Spertus, J. A., Lindau, S. T., Morgan, T. M., Herrin, J., Bueno, H., Mattera, J. A., Ridker, P. M., & Krumholz, H. M. (2010). Variation in recovery: Role of gender on outcomes of young AMI patients (VIRGO) study design. Circulation. Cardiovascular Quality and Outcomes, 3(6), 684–693. https://doi.org/10.1161/CIRCOUTCOMES.109.928713

14. ^14^Vaccarino, V., Parsons, L., Every, N. R., Barron, H. V., & Krumholz, H. M. (1999). Sex-based differences in early mortality after myocardial infarction. National Registry of Myocardial Infarction 2 Participants. The New England journal of medicine, 341(4), 217–225. https://doi.org/10.1056/NEJM199907223410401

15. Grundy, S. M., Stone, N. J., Bailey, A. L., Beam, C., Birtcher, K. K., Blumenthal, R. S., Braun, L. T., de Ferranti, S., Faiella-Tommasino, J., Forman, D. E., Goldberg, R., Heidenreich, P. A., Hlatky, M. A., Jones, D. W., Lloyd-Jones, D., Lopez-Pajares, N., Ndumele, C. E., Orringer, C. E., Peralta, C. A., Saseen, J. J., … Yeboah, J. (2019). 2018 AHA/ACC/AACVPR/AAPA/ABC/ACPM/ADA/AGS/AphA/ASPC/NLA/PCNA Guideline on the Management of Blood Cholesterol: A Report of the American College of Cardiology/American Heart Association Task Force on Clinical Practice Guidelines. Circulation, 139(25), e1082–e1143. https://doi.org/10.1161/CIR.0000000000000625

16. Pearson, G. J., Thanassoulis, G., Anderson, T. J., Barry, A. R., Couture, P., Dayan, N., Francis, G. A., Genest, J., Grégoire, J., Grover, S. A., Gupta, M., Hegele, R. A., Lau, D., Leiter, L. A., Leung, A. A., Lonn, E., Mancini, G., Manjoo, P., McPherson, R., Ngui, D., … Wray, W. (2021). 2021 Canadian Cardiovascular Society Guidelines for the Management of Dyslipidemia for the Prevention of Cardiovascular Disease in Adults. The Canadian Journal of Cardiology, 37(8), 1129–1150. https://doi.org/10.1016/j.cjca.2021.03.016

17. Wenger, N. K., Lloyd-Jones, D. M., Elkind, M., Fonarow, G. C., Warner, J. J., Alger, H. M., Cheng, S., Kinzy, C., Hall, J. L., Roger, V. L., & American Heart Association (2022). Call to Action for Cardiovascular Disease in Women: Epidemiology, Awareness, Access, and Delivery of Equitable Health Care: A Presidential Advisory From the American Heart Association. Circulation, 101161CIR0000000000001071. Advance online publication. https://doi.org/10.1161/CIR.0000000000001071

18. Safdar, B., Spatz, E. S., Dreyer, R. P., Beltrame, J. F., Lichtman, J. H., Spertus, J. A., Reynolds, H. R., Geda, M., Bueno, H., Dziura, J. D., Krumholz, H. M., & D’Onofrio, G. (2018). Presentation, Clinical Profile, and Prognosis of Young Patients With Myocardial Infarction With Nonobstructive Coronary Arteries (MINOCA): Results From the VIRGO Study. Journal of the American Heart Association, 7(13), e009174. https://doi.org/10.1161/JAHA.118.009174

19. Saw, J., Humphries, K., Aymong, E., Sedlak, T., Prakash, R., Starovoytov, A., & Mancini, G. B. J. (2017). Spontaneous Coronary Artery Dissection: Clinical Outcomes and Risk of Recurrence. Journal of the American College of Cardiology, 70(9), 1148–1158. https://doi.org/10.1016/j.jacc.2017.06.053

20. ^20^Hayes, S. N., Kim, E. S. H., Saw, J., Adlam, D., Arslanian-Engoren, C., Economy, K. E., Ganesh, S. K., Gulati, R., Lindsay, M. E., Mieres, J. H., Naderi, S., Shah, S., Thaler, D. E., Tweet, M. S., Wood, M. J., & American Heart Association Council on Peripheral Vascular Disease; Council on Clinical Cardiology; Council on Cardiovascular and Stroke Nursing; Council on Genomic and Precision Medicine; and Stroke Council (2018). Spontaneous Coronary Artery Dissection: Current State of the Science: A Scientific Statement From the American Heart Association. Circulation, 137(19), e523–e557. https://doi.org/10.1161/CIR.0000000000000564

21. ^21^Tamis-Holland, J. E., Jneid, H., Reynolds, H. R., Agewall, S., Brilakis, E. S., Brown, T. M., Lerman, A., Cushman, M., Kumbhani, D. J., Arslanian-Engoren, C., Bolger, A. F., Beltrame, J. F., & American Heart Association Interventional Cardiovascular Care Committee of the Council on Clinical Cardiology; Council on Cardiovascular and Stroke Nursing; Council on Epidemiology and Prevention; and Council on Quality of Care and Outcomes Research (2019). Contemporary Diagnosis and Management of Patients With Myocardial Infarction in the Absence of Obstructive Coronary Artery Disease: A Scientific Statement From the American Heart Association. Circulation, 139(18), e891–e908. https://doi.org/10.1161/CIR.0000000000000670

22. ^22^Kunadian, V., Chieffo, A., Camici, P. G., Berry, C., Escaned, J., Maas, A. H. E. M., Prescott, E., Karam, N., Appelman, Y., Fraccaro, C., Louise Buchanan, G., Manzo-Silberman, S., Al-Lamee, R., Regar, E., Lansky, A., Abbott, J. D., Badimon, L., Duncker, D. J., Mehran, R., Capodanno, D., … Baumbach, A. (2020). An EAPCI Expert Consensus Document on Ischaemia with Non-Obstructive Coronary Arteries in Collaboration with European Society of Cardiology Working Group on Coronary Pathophysiology & Microcirculation Endorsed by Coronary Vasomotor Disorders International Study Group. European heart journal, 41(37), 3504–3520. https://doi.org/10.1093/eurheartj/ehaa503

23. ^23^Ong, P., Camici, P. G., Beltrame, J. F., Crea, F., Shimokawa, H., Sechtem, U., Kaski, J. C., Bairey Merz, C. N., & Coronary Vasomotion Disorders International Study Group (COVADIS) (2018). International standardization of diagnostic criteria for microvascular angina. International journal of cardiology, 250, 16–20. https://doi.org/10.1016/j.ijcard.2017.08.068

24. ^24^Brunham, L. R., Lynch, K., English, A., Sutherland, R., Weng, J., Cho, R., Wong, G. C., Anis, A. H., Francis, G. A., Khan, N. A., McManus, B., Wood, D., Walley, K. R., Leipsic, J., Humphries, K. H., Hoens, A., Krahn, A. D., John Mancini, G. B., & Pimstone, S. (2018). The design and rationale of SAVE BC: The Study to Avoid CardioVascular Events in British Columbia. Clinical Cardiology, 41(7), 888–895. https://doi.org/10.1002/clc.22959

25. National Cholesterol Education Program (NCEP) Expert Panel on Detection, Evaluation, and Treatment of High Blood Cholesterol in Adults (Adult Treatment Panel III) (2002). Third Report of the National Cholesterol Education Program (NCEP) Expert Panel on Detection, Evaluation, and Treatment of High Blood Cholesterol in Adults (Adult Treatment Panel III) final report. Circulation, 106(25), 3143–3421.

26. Rabi, D. M., McBrien, K. A., Sapir-Pichhadze, R., Nakhla, M., Ahmed, S. B., Dumanski, S. M., Butalia, S., Leung, A. A., Harris, K. C., Cloutier, L., Zarnke, K. B., Ruzicka, M., Hiremath, S., Feldman, R. D., Tobe, S. W., Campbell, T. S., Bacon, S. L., Nerenberg, K. A., Dresser, G. K., Fournier, A., … Daskalopoulou, S. S. (2020). Hypertension Canada’s 2020 Comprehensive Guidelines for the Prevention, Diagnosis, Risk Assessment, and Treatment of Hypertension in Adults and Children. The Canadian journal of cardiology, 36(5), 596–624. https://doi.org/10.1016/j.cjca.2020.02.086

27. Diabetes Canada Clinical Practice Guidelines Expert Committee, Punthakee, Z., Goldenberg, R., & Katz, P. (2018). Definition, Classification and Diagnosis of Diabetes, Prediabetes and Metabolic Syndrome. Canadian journal of diabetes, 42 *Suppl 1*, S10–S15. https://doi.org/10.1016/j.jcjd.2017.10.003

28. Gestational Hypertension and Preeclampsia: ACOG Practice Bulletin, Number 222. (2020). Obstetrics and gynecology, 135(6), e237–e260. https://doi.org/10.1097/AOG.0000000000003891

29. Nakamura, Y., Shinozaki, N., Hirasawa, M., Kato, R., Shiraishi, K., Kida, H., Usuda, K., & Ishikawa, T. (2000). Prevalence of migraine and Raynaud’s phenomenon in Japanese patients with vasospastic angina. Japanese circulation journal, 64(4), 239–242. https://doi.org/10.1253/jcj.64.239

30. Godo, S., Suda, A., Takahashi, J., Yasuda, S., & Shimokawa, H. (2021). Coronary Microvascular Dysfunction. Arteriosclerosis, thrombosis, and vascular biology, 41(5), 1625– 1637. https://doi.org/10.1161/ATVBAHA.121.316025

31. Bastiany, A., Pacheco, C., Sedlak, T., Saw, J., Miner, S. E. S., Liu, S., Lavoie, A., Kim, D. H., Gulati, M., & Graham, M. M. (2022). A Practical Approach to Invasive Testing in Ischemia With No Obstructive Coronary Arteries (INOCA). CJC open, 4(8), 709–720. https://doi.org/10.1016/j.cjco.2022.04.009

32. Honigberg, M. C., Ye, Y., Dattilo, L., Sarma, A. A., Scott, N. S., Smoller, J. W., Zhao, H., Wood, M. J., & Natarajan, P. (2022). Low depression frequency is associated with decreased risk of cardiometabolic disease. Nature Cardiovascular Research. 1: 125–131. https://doi.org/10.1038/s44161-021-00011-7

33. Mehta, P. K., Thobani, A., & Vaccarino, V. (2019). Coronary Artery Spasm, Coronary Reactivity, and Their Psychological Context. Psychosomatic medicine, 81(3), 233–236. https://doi.org/10.1097/PSY.0000000000000682

34. Goncharova N. D. (2013). Stress responsiveness of the hypothalamic-pituitary-adrenal axis: age-related features of the vasopressinergic regulation. Frontiers in endocrinology, 4, 26. https://doi.org/10.3389/fendo.2013.00026

35. ^35^Kinder, J. M., Stelzer, I. A., Arck, P. C., & Way, S. S. (2017). Immunological implications of pregnancy-induced microchimerism. Nature reviews. Immunology, 17(8), 483–494. https://doi.org/10.1038/nri.2017.38

36. Lv, H., Wu, H., Yin, J., Qian, J., & Ge, J. (2015). Parity and Cardiovascular Disease Mortality: a Dose-Response Meta-Analysis of Cohort Studies. Scientific reports, 5, 13411. https://doi.org/10.1038/srep13411

37. Saarelainen, H., Valtonen, P., Punnonen, K., Laitinen, T., Raitakari, O. T., Juonala, M., Heiskanen, N., Lyyra-Laitinen, T., Viikari, J. S., & Heinonen, S. (2009). Flow mediated vasodilation and circulating concentrations of high sensitive C-reactive protein, interleukin-6 and tumor necrosis factor-alpha in normal pregnancy—The Cardiovascular Risk in Young Finns Study. Clinical physiology and functional imaging, 29(5), 347–352. https://doi.org/10.1111/j.1475-097X.2009.00877.x

38. Jacobs, M. B., Kritz-Silverstein, D., Wingard, D. L., & Barrett-Connor, E. (2012). The association of reproductive history with all-cause and cardiovascular mortality in older women: the Rancho Bernardo Study. Fertility and sterility, 97(1), 118–124. https://doi.org/10.1016/j.fertnstert.2011.10.028

39. Parikh, N. I., Cnattingius, S., Dickman, P. W., Mittleman, M. A., Ludvigsson, J. F., & Ingelsson, E. (2010). Parity and risk of later-life maternal cardiovascular disease. American heart journal, 159(2), 215–221.e6. https://doi.org/10.1016/j.ahj.2009.11.017

40. Joffres, M., Shields, M., Tremblay, M. S., & Connor Gorber, S. (2013). Dyslipidemia prevalence, treatment, control, and awareness in the Canadian Health Measures Survey. Canadian journal of public health, 104(3), e252–e257. https://doi.org/10.17269/cjph.104.3783

41. Public Health Agency of Canada (2019). Twenty Years of Diabetes surveillance using the Canadian Chronic Disease Surveillance System. Available from: https://www.canada.ca/content/dam/phac-aspc/documents/services/publications/diseases-conditions/twenty-years-of-diabetes/64-03-19-2467-Diabetes-Infographic-EN-11.pdf

42. Government of Canada (2022). Canadian Tobacco and Nicotine Survey (CTNS): Summary of Results for 2020. Available from: https://www.canada.ca/en/health-canada/services/31anadian-tobacco-nicotine-survey/2020-summary.html

43. Ramage-Morin, P.L. & Gilmour, H. (2014). Prevalence of migraine in the Canadian household population. Components of Statistics Canada Catalogue no. 82-003-X: Health Reports. Accessed from: https://www150.statcan.gc.ca/n1/pub/82-003-x/2014006/article/14033-eng.pdf

44. Sueda, S., Kohno, H., Fukuda, H., Ochi, N., Kawada, H., Hayashi, Y., & Uraoka, T. (2004). Frequency of provoked coronary spasms in patients undergoing coronary arteriography using a spasm provocation test via intracoronary administration of ergonovine. Angiology, 55(4), 403–411. https://doi.org/10.1177/000331970405500407

45. Hung, M. Y., Hsu, K. H., Hung, M. J., Cheng, C. W., & Cherng, W. J. (2010). Interactions among gender, age, hypertension and C-reactive protein in coronary vasospasm. European journal of clinical investigation, 40(12), 1094–1103. https://doi.org/10.1111/j.1365-2362.2010.02360.x

46. Shin, D. I., Baek, S. H., Her, S. H., Han, S. H., Ahn, Y., Park, K. H., Kim, D. S., Yang, T. H., Choi, D. J., Suh, J. W., Kwon, H. M., Lee, B. K., Gwon, H. C., Rha, S. W., & Jo, S. H. (2015). The 24-Month Prognosis of Patients With Positive or Intermediate Results in the Intracoronary Ergonovine Provocation Test. JACC. Cardiovascular interventions, 8(7), 914–923. https://doi.org/10.1016/j.jcin.2014.12.249

